# Predicting the Progression of Mild Cognitive Impairment Using Machine Learning: A Systematic, Quantitative and Critical Review

**DOI:** 10.1101/2020.09.01.20185959

**Authors:** Manon Ansart, Stéphane Epelbaum, Giulia Bassignana, Alexandre Bône, Simona Bottani, Tiziana Cattai, Raphaël Couronné, Johann Faouzi, Igor Koval, Maxime Louis, Elina Thibeau-Sutre, Junhao Wen, Adam Wild, Ninon Burgos, Didier Dormont, Olivier Colliot, Stanley Durrleman

## Abstract

We performed a systematic review of studies focusing on the automatic prediction of the progression of mild cognitive impairment to Alzheimer’s disease (AD) dementia, and a quantitative analysis of the methodological choices impacting performance. This review included 172 articles, from which 234 experiments were extracted. For each of them, we reported the used data set, the feature types, the algorithm type, performance and potential methodological issues. The impact of these characteristics on the performance was evaluated using a multivariate mixed effect linear regressions. We found that using cognitive, fluorodeoxyglucose-positron emission tomography or electroencephalography and magnetoencephalography variables significantly improved predictive performance compared to not including them, whereas including other modalities, in particular T1 magnetic resonance imaging, did not show a significant effect. The good performance of cognitive assessments questions the wide use of imaging for predicting the progression to AD and advocates for exploring further fine domain-specific cognitive assessments. We also identified several methodological issues, including the absence of a test set, or its use for feature selection or parameter tuning in nearly a fourth of the papers. Other issues, found in 15% of the studies, cast doubts on the relevance of the method to clinical practice. We also highlight that shortterm predictions are likely not to be better than predicting that subjects stay stable over time. These issues highlight the importance of adhering to good practices for the use of machine learning as a decision support system for the clinical practice.

## 1. Introduction

The early diagnosis of Alzheimer’s disease (AD) is crucial for patient care and treatment. Machine learning algorithms have been used to perform automatic diagnosis and predict the current clinical status at an individual level, mainly in research cohorts. Individuals suffering from mild cognitive impairment (MCI) are however likely to have a change of clinical status in the coming years, and to be diagnosed with AD or another form of dementia. Distinguishing between the MCI individuals that will remain MCI (MCI stable, or sMCI) from those who will progress to AD (pMCI) is an important task, that can allow for the early care and treatment of pMCI patients. In this article, we will review methods that have been proposed to automatically predict if an MCI patient will develop AD dementia in the future by performing a careful reading of published articles, and compare them through a quantitative analysis.

The application of machine learning to precision medicine is an emerging field, at the cross roads of different disciplines, such as computer science, radiology or neurology. Researchers working on the topic usually come from one field or the other, and therefore do not have all the skills that are necessary to design methods that would be efficient and following machine learning best practices, while being understandable and useful to clinicians.

Reviews of the automatic prediction of the patient’s current diagnosis from clinical or imaging variables acquired at the same time in the context of AD have already been published, but none specifically target the prediction of progression from MCI to AD dementia. They focus on the use of magnetic resonance imaging (MRI) (Falahati et al., 2014; Leandrou et al., 2018), or of neuroimaging data more broadly (Rathore et al., 2017; Arbabshirani et al., 2017; Haller et al., 2011; Sarica et al., 2017). Several of them are systematic reviews such as Arbabshirani et al. (2017) with 112 studies on AD, Rathore et al. (2017) with 81 studies, Falahati et al. (2014) with 50 studies and Sarica et al. (2017) with 12 studies. They often gather the findings of each individual article and compare them, but no quantitative analysis of performance is proposed.

We propose here to perform a systematic and quantitative review of studies predicting the evolution of clinical diagnosis in individuals with MCI. We will report different quantitative and qualitative characteristics of the proposed method such as the sample size, type of algorithm, reported accuracy, identification of possible issues. We will then analyze this data to identify the characteristics which impact performance the most, and list several recommendations to ensure that the performance is well estimated, and that the algorithm would have the best chance to be useful in clinical practice.

## 2. Materials and Method

### 2.1. Selection process

The query used to find the relevant articles was composed of 4 parts:

1. As we study the progression from MCI to AD, the words MCI and AD should be present in the abstract;
2. We removed the articles predicting only the patient’s current diagnosis using variables acquired at the same time point by ensuring the words “prediction” and “progression” or associated terms are present in the abstract;
3. A performance measure should be mentioned;
4. A machine learning algorithm or classification related key-word should be in the abstract. This fourth part ensures the selected articles make individual predictions and reduces the presence of group analyses.

The full query can be found in Appendix A.1. Running it on Scopus on the *13^th^* of December 2018 resulted in 330 articles. The abstracts were read to remove irrelevant articles, including studies of the progression of cognitively normal individuals to MCI, automatic diagnosis methods, review articles and group analyses. After this selection 206 articles were identified. As this first selection was quite conservative, 34 additional articles were removed from the selection for similar reasons during the reading process, leaving 172 studied articles which are listed in Appendix B. The selection process is described in Figure S1 in Appendix A.2.

### 2.2. Reading process

For each study, the number of individuals was first assessed and noted. Only studies including more than 30 sMCI and 30 pMCI (111 articles) were then fully read, as we considered that experience using fewer than 30 individuals cannot provide robust estimates of performance. Articles with fewer than 30 individuals in each category were still considered when studying the evolution of the number of articles with time, and of the number of individuals per article with time. The studies including enough individuals were then analyzed by one of the 19 readers participating in this review, and a final curation was performed by one of the authors (MA) to ensure homogeneity. 36 items, of which a list is available in Appendix A.3, were reported for each study, including the used features, the cohort, the method (time to prediction, algorithm, feature selection, feature processing), the evaluation framework and the performance measures, as well as identified biases in the method. When several experiments were available in an article, they were all reported in the table. A total of 234 experiments was thus studied.

A table containing the articles included in the review and the reported values can be found on https://gitlab.com/icm-institute/aramislab/mci-progression-review. The issues identified in each article were removed from this open-access table, to avoid negatively pointing at studies. They can be made available if requested to the corresponding author.

### 2.3. Quality check

Several methodological issues were identified during the reading process. This list of issues was not previously defined, it has been established as issues were encountered in the various studies. We identified the following list of issues:

- Lack of a test data set: use of the same data set for training and testing the algorithm, without splitting the data set or using any kind of cross-validation method. The performance computed this way is the training performance, whereas a test performance, computed on a different set of individuals, is necessary to measure the performance that could be obtained on another data set (i.e. generalizability of the method).
- Automatic feature selection performed on the whole data set. When a large number of features is available, automatic feature selection can be performed in order to identify the most relevant features and use them as input. A variety of automatic algorithms exist to do this. Some studies performed this automatic feature selection on the whole data set, before splitting it into a training and a test set or performing cross-validation. An example of this issue is, first, using t-tests to identify features that best separate pMCI from sMCI, using the whole data set, then splitting the data set into a training and a test set, to respectively train the classification algorithm and evaluate its performance. In this example, the individuals from the test set have been used to perform the automatic feature selection and choose the most relevant features. This is an issue, as individuals in the test set should be used for performance evaluation only.
- Other data-leakage. More broadly, data leakage is the use of data from the test set outside of performance evaluation. Using the test data set for parameter tuning, or for choosing the best data set out of a large number of experiments, are two common examples of data leakage.
- Feature embedding performed on the whole data set. Feature embedding (for example principal components analysis) transforms the input features into a lower-dimension feature space. It is often used to reduce the input dimension when many features are available, but it does not use the individual labels (sMCI/pMCI) to do so, as feature selection often does. This issue is therefore similar to performing feature selection on the whole data set, except that only the features of the test individuals are used, and not their labels.
- Use of the date of AD diagnosis to select the input visit of pMCI individuals. An example of this issue is using the visit 3 years before progression to AD for pMCI subjects, and the first available visit for sMCI subjects, to predict the progression to AD at 3 years, even for testing the method. In this case, the date of progression to AD of the individuals of the test set was used to select the input visit, which is not possible in clinical practice, as the date of progression is not known. Such experimental designs are also likely to introduce biases between pMCI and sMCI subjects in age or in total observation periods for instance, which may lead to a better performance than what could be achieved in a real-life scenario.

Other methodological issues, not belonging to these categories, were also reported, such as incompatibility between different reported measures. The articles in which at least one of these issues was identified were not used when analyzing the performance of the methods. Only articles with no reported issues were used, however it is possible that some issues could not be detected from the elements given in the articles, and that some issues were not identified during reading.

### 2.4. Statistical analysis

#### 2.4.1. General model

We studied the impact of various method characteristics (such as input feature and algorithm) on the performance of the classification task, separating sMCI from pMCI individuals. Several experiments were reported for each article, so we had to account for the dependency between experiments coming from the same article. In order to do so, we used linear mixed-effects models with a random intercept on the article.

For the model to have enough power, we grouped the characteristics in a hierarchical manner, creating broad categories that can be expanded into finer ones several times. The categories were created as such:

- linear models: linear regression, orthogonal partial least square (OPLS), linear discriminant analysis (LDA), manual threshold
- generalized linear models: linear support vector machine(SVM), logistic regression, survival analysis
- non-linear models: random forest, multi-kernel learning, non-linear SVM, Bayesian methods, neural networks, others
- imaging features

– T1 MRI

- region-based features on selected regions of interest (T1-ROI)
- region-based features on the whole brain
- voxel-based features
– positron emission tomography (PET)

- fluorodeoxyglucose (FDG) PET
- Amyloid PET
– white matter hyper-intensities
– electroencephalography (EEG) or magnetoencephalography (MEG)
– diffusion tensor imaging (DTI)
– fMRI
• cerebrospinal fluid (CSF) biomarkers
• cognitive features
– general cognitive features
– domain-targeted cognitive features
– new, home-made cognitive features
• socio-demographic and genetic features
– socio-demographic features

- age
- gender
– Apolipoprotein E (APOE)
- other features
- longitudinal approach
- use of the Alzheimer’s Disease Neuroimaging Initiative (ADNI) data set
- number of subjects

A first model was created with the broadest categories, and we used a two-sided t-test on the regression coefficients to identify the categories of characteristics which had a significant impact on performance. The next model was then created by expanding only the significant categories and keeping the non-significant one at a coarse level.

The expansion and the creation of new models was repeated until we reached a model for which all significant coefficients belonged to categories that could not be expanded further. We report the results of the final model in 4. The intermediate models leading to the final one are reported in section Appendix A.4.1 of Supplementary Materials.

For each model, we only used the characteristics which were found in more than one article with an associated performance measure and with no identified issue. The performance measure used for these models was the area under the receiver operating characteristic (ROC) curve (AUC), experiments with no reported AUC were therefore not taken into account.

Only the experiments with no identified methodological issues were included in the model. This process was performed twice: once using all experiments without issues, and once using only the experiments performed on the ADNI database.

The p-values corrected for multiple comparisons were obtained by using the Benjamini-Hochberg procedure.

#### 2.4.2. Individual feature models

We wanted to test whether T1 MRI, cognitive or FDG PET features are predictors of better performance if used alone or in combination with other features. To this purpose, for a given feature type *F*, we selected the experiments using this feature type and that had a reported AUC and no methodological issue. We then used a linear mixed-effect model, defined as:

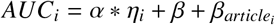

where i is the experiment, *article_i_* is the article to which the experiment belongs (as several experiments can be reported in each article), and *η_i_* is 0 when the experiment uses only the studied feature type *F* and 1 when it uses other feature types as well. We used a two-sided t-test on *α* to determine if including other feature types significantly changed the performance compared to using *F* alone.

This analysis was performed for F being: (a) T1 MRI features, (b) cognitive features, (c) FDG PET features. These were the features selected for the final general model as explained in 2.4.1, that have been used alone in at least 2 reported experiments, and that have been used in combination with other features in at least 2 experiments as well. Cognitive features were not divided into subsets so as to study the effect of cognitive assessments as a whole.

## 3. Descriptive analysis

### 3.1. A recent trend

Figure 1a shows that the number of articles published each year on the prediction of the progression of MCI to AD dementia has been steadily increasing since 2010.

**Figure 1:**
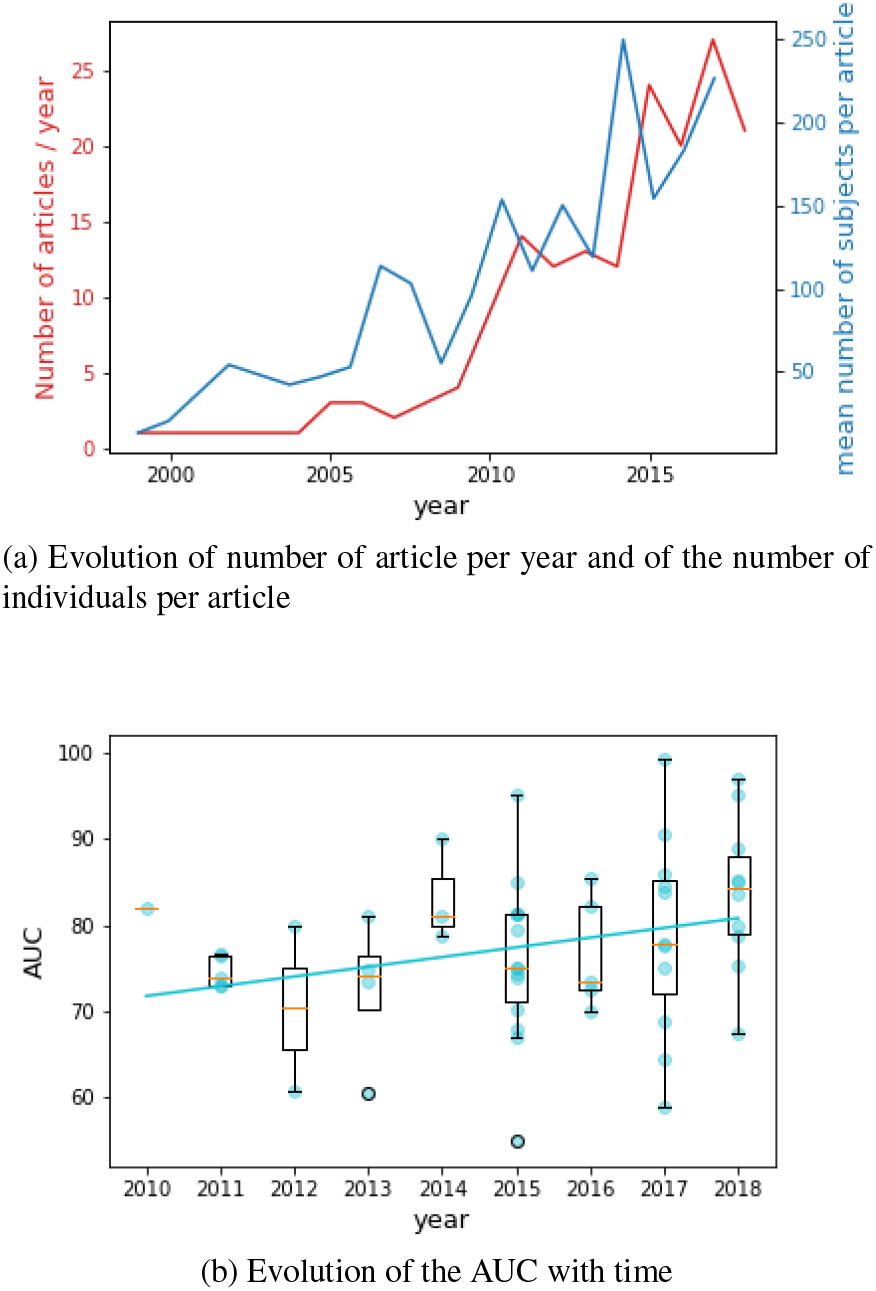
Recent trends. (a) Evolution of number of article per year (in red) and of the number of individuals per article with time (in blue). (b) Evolution of the area under the ROC (receiver operating characteristic) curve (AUC) with time. The AUC of each article is represented by a dot. The AUC of articles published the same year is represented as box-plots. The plain line corresponds to the regression of the AUC against time

Figure 1a also shows that the number of individuals used for the experiments is increasing over time (p= 10−^5^, slope of 12.15 subjects per year, *R*^2^ *=* 0.10). 84.6% of articles used data of the Alzheimer’s Disease Neuroimaging Initiative (ADNI) study. Starting in 2004, this multicenter longitudinal study provides multiple modalities for the early detection of AD. As the recruitment of this largely used cohort is still ongoing, it is not surprising to see the number of included individuals increasing over the years. Studies often select individuals with a minimal follow-up time, of 3 years for example, and over the years more and more MCI individuals from the ADNI cohort fulfill these criteria, so more individuals can be included.

As shown in Figure 1b, the reported AUC are also increasing over time (p= 0.045, slope of 1.15 points of AUC per year), which can have multiple explanations. First, as new studies often compare their performance with those of previous methods, they tend to be published only when the obtained results seem competitive compared to previous ones. A more optimistic interpretation would be that algorithms tend to improve, and that newly available features might have a better predictive power. It has also been shown (Ansart et al., 2019; Domingos, 2012) that having a larger data set leads to a higher performance, so there may be a link between the increase in data set size and the increase in performance.

### 3.2. Features

T1 MRI, cognition and socio-demographic features are used in respectively 69.2%, 43.2% and 33.8% of experiments. On the other hand, FDG PET, APOE and CSF AD biomarkers are used in 15 to 20% of experiments, and the other studied features (white matter hyper-intensities, EEG, MEG, PET amyloid, amyloid binary status without considering the PET or CSF value, DTI and PET Tau) are used in fewer than 10% of experiments. No study using functional MRI has been identified.

Studies using T1 MRI mainly use selected regions of interest (46.8%), whereas 34.7% use the whole brain, separated into regions of interest, and 18.5% use voxel features. Studies using neuro-psychological tests mainly use aggregated tests evaluating multiple domains of cognition (51.2% of them), and 37.4% of them combine aggregated tests with domain-specific ones. Seven experiments use new or home-made cognitive tests. 35.7% of experiments use only T1 MRI (apart from socio-demographic features), and 15% use cognition only.

The prevalence of T1 MRI does not seem surprising, as researchers working on automatic diagnosis often come from the medical imaging community, and T1 MRI is the most widely available modality. The prevalence of the imaging community can also explain the choice of cognitive features, and why more detailed and targeted cognitive tests are not used as much as more general and more well-known ones.

### 3.3. Algorithm

Support vector machines (SVM) and logistic regressions are the most used algorithms, being used in respectively 32.6% and 15.0% of experiments. Among the experiments using an SVM, 63.2% use a non-linear kernel, 30.3% use a linear kernel and 6.6% do not mention the used kernel. Other algorithms are used in fewer than 10% of cases. Figure 2 shows the evolution of the algorithm use over time.

**Figure 2:**
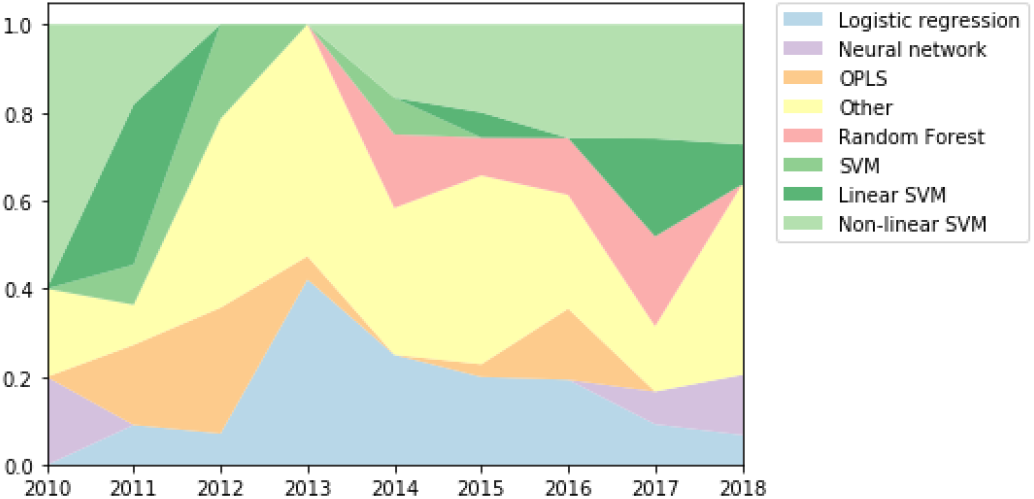
Evolution of the use of various algorithms with time. SVM with unknown kernel are simply noted as "SVM". OPLS: orthogonal partial least square; SVM: support vector machine

The high proportion of methods using an SVM has already been shown for the prediction of the current diagnosis in Falahati et al. (2014) and Rathore et al. (2017), it is therefore not surprising that this algorithm is also commonly used for the prediction of future diagnosis. We see that random forests started being used around 2014, but the proportion of methods using this algorithm, even recently, stays low compared to the proportion of methods using an SVM. Neural networks started being used during the last two years, as it can be seen in Figure 2, and we can assume the phenomenon has been too recent to be visible just yet in the field. Overall, even if the proportion of SVM has been decreasing until 2013, the field has not been so prompt to use new algorithms as one could have expected. A possible explanation is that the choice of algorithm does not significantly impact performance.

### 3.4. Validation method

For evaluating their performance, 29.1 % of experiments use a 10-fold, and 12.8% use a k-fold with k different from 10. Leave-one individual out is also quite popular, being used in 17.5% of cases. We noted that 7.3% of experiments were trained and tested on the same individuals, and 7.3% train the method on a first cohort and test it on a different one.

It should be kept in mind when comparing the performance of different studies that the cross-validation methods can impact the performance. Using a larger training set and smaller test set is more favorable, hence the same method might result in a better performance when evaluated using a leave-one out validation than using a 10-fold validation, as shown in Lin et al. (2018). Bias and variance also vary across validation methods (Efron, 1983). Varoquaux et al. (2017) also studied the impact of the crossvalidation strategy on a range of classification tasks performed on fMRI and MEG data sets, and showed that differences in performance tend to be smaller than the variance of the estimated performance using a cross-validation method, mitigating the importance of the choice of cross-validation strategy. This study still warns against the use of leave-one-out validation, leading to less stable estimates.

Reporting variance or confidence intervals is an important best practice to compare results from different studies and experiments. We did not collect this information, and further work regarding the adoption of this practice could complete this study.

## 4. Performance analyses

The results of the linear mixed-effect model used to model the AUC based on the method characteristics are shown in Table 1, and the details of the intermediate models can be found in section Appendix A.4.1 of Supplementary Materials. The performance is significantly better when using EEG and MEG (coefficient=3.4, p=3*10^-3^), domain-targeted cognitive features (coefficient=2.6, p = 0.026), FDG PET (coefficient=2.2, p=0.023) or APOE (coefficient=2.3, p=0.049). The use of the ADNI cohort and of longitudinal data are not shown to be significant. The impact of the algorithm type and of the number of subjects are not shown to be significant either.

**Table 1:** Impact of method characteristics. This table shows the coefficients obtained using the linear mixed- effect model described in section 2.4.1 on all experiments, the associated p-values and corrected p-values. The last columns shows the number of experiments using the given characteristic, out of the 120 experiments included in the model. Benjamini-Hochberg procedure was applied to get corrected p-values. coeff.:coefficient of the characteristics in the mixed effect model; FDG: fluorodeoxyglucose; PET: positron emission tomography; EEG: electroencephalography; MEG: magnetoencephalography; APOE: Apolipopro- tein E; ADNI: Alzheimer’s Disease Neuroimaging Initiative; NA: not applicable

| Characteristic | coeff. | p-value | corrected p-value | number of exp. |
| --- | --- | --- | --- | --- |
| intercept | 78 | **0** | **0** | NA |
| linear model | -0.47 | 0.79 | 0.94 | 23 |
| generalized linear model | 0.19 | 0.89 | 0.96 | 28 |
| non linear model | 0.83 | 0.55 | 0.79 | 50 |
| T1 features | 0.92 | 0.26 | 0.76 | 77 |
| amyloid PET | 1.3 | 0.35 | 0.79 | 5 |
| FDG PET | 2.6 | **0.023** | 0.13 | 24 |
| white matter hyper-intensities | -0.58 | 0.49 | 0.79 | 3 |
| EEG/MEG | 3.4 | **$2.9 \times 10^{-03}$** | **0.029** | 5 |
| general cognitive features | -0.14 | 0.91 | 0.96 | 49 |
| domain targeted cognitive features | 2.6 | **0.026** | 0.13 | 25 |
| new or specific cognitive features | 0.89 | 0.52 | 0.79 | 2 |
| socio-demographic features | 1.2 | 0.43 | 0.79 | 43 |
| APOE | 2.27 | **0.049** | 0.19 | 26 |
| biomarkers | 0.75 | 0.39 | 0.79 | 19 |
| other features | 0.53 | 0.54 | 0.79 | 12 |
| longitudinal | 0.25 | 0.80 | 0.95 | 13 |
| ADNI | 0.011 | 0.99 | 0.99 | 106 |
| number of subjects | -0.39 | 0.76 | 0.94 | NA |
| individual intercept | NA | 0.072 | 0.24 | NA |

We also run the performance analysis using only the experiments performed on the ADNI cohort. The only characteristics with a significant impact on the AUC are the use of T1-ROI features (coefficient = 1.7, p=0.014) (and not the other T1-based features which are regions based features on the whole brain and voxel-based features), FDG PET features (coefficient = 4.4, p < 1 * 10^-7^) and domain-targeted cognitive features (coefficient=2.4, p = 9*10^-3^). The complete results can be found in section Appendix A.4.2 of Supplementary Materials.

We considered the impact of using each feature alone compared to a combination of them. It is significantly better to combine T1 MRI with other features than to use it solely (coefficient = 5.5, p = 9*10^-3^). The effect is not significant for cognition (coefficient=3.0, p=0.19) and FDG PET (coefficient = -6.1, p=0.38).

### 4.1. Cognition

Cognitive variables can be easily collected in clinical routine, at a low cost, and they are proven to increase the performance of the methods, so their use should be encouraged. This finding is consistent with comparisons performed in several studies. Minhas et al. (2018); Kauppi et al. (2018); Ardekani et al. (2017); Tong et al. (2017); Gavidia-Bovadilla et al. (2017); Moradi et al. (2015); Hall et al. (2015); Fleisher et al. (2008) showed that using cognition and T1 MRI performed better than using T1 MRI only. Dukart et al. (2015); Cui et al. (2011); Thung et al. (2018); Li et al. (2018) showed that adding cognition to other modalities also improved the results.

More surprisingly, we showed that using other modalities does not significantly improve the results compared to using cognition only. Although Fleisher et al. (2008) shows that using T1 MRI in addition to cognition does not improve the performance compared to using cognition only, several studies show the opposite on various modalities (Samper-Gonzalez et al., 2019; Moradi et al., 2015; Ardekani et al., 2017; Li et al., 2018; Kauppi et al., 2018). However, when taking all studies into account, it appears that the improvement one gains by including other modalities along with cognitive variables is not significant. As the cost of collecting cognitive variables compared to performing an MRI or a FDG PET is quite low, the non-significant improvement in performance might not be worth the cost, logistics and patient inconvenience arising from the collection of other modalities. Methods focusing on cognition only, such as proposed by Johnson et al. (2014), should therefore be further explored. Such methods should include domain-specific cognitive scores, which have shown to increase the performance.

### 4.2. Medical imaging and biomarkers

Imaging modalities are not as widely available as cognitive features, but they can represent a good opportunity to better understand the disease process by showing the changes that appear before the individuals progress to AD dementia.

Among the used imaging modalities, we showed that using FDG PET leads to a better performance. Using T1-ROI features also leads to a better performance on the ADNI experiments, but this effect is not significant when considering all experiments. All the experiments using T1 MRI are performed on the ADNI database, so one can assume the effect on performance is small and is diluted when considering all experiments instead of the experiments performed on ADNI. Even considering the ADNI experiments, the effect of using T1-ROI features is 2.6 times smaller than the effect of using FDG PET, and 1.45 times smaller than the effect of using cognitive features. We also showed that using T1 MRI features alone performs significantly worse than using other features as well. Over all, T1 MRI features should not be used alone and PET images could represent a better alternative for the imaging community. Similar observations have been made by Samper-Gonzalez et al. (2018). FDG PET was included as a supportive feature in AD diagnosis criteria in 2007 (Dubois et al., 2007), and although it was removed - along with structural MRI - from IWG-2 diagnostic criteria in 2014, Dubois et al. (2014) stressed that FDG PET can be useful to differentiate between AD and other types of dementia and to measure disease progression. According to the model hypothesized in (Jack et al., 2010a) changes in FDG PET appear earlier in the AD process than changes in structural MRI, which has been corroborated by different quantitative studies (Chetelat et al., 2007; Reiman et al., 1998; Jagust et al., 2006). These changes might already be visible in MCI individuals several years before their progression to AD, which can explain why FDG PET is more predictive of this progression.

Only one method using Tau PET has been identified in this review so we could not evaluate the impact on performance. This new modality should also be affected early in the disease process, and could therefore represent great hopes for the imaging community. However, surprisingly, Amyloid PET or CSF value, which is also one of the earliest markers, did not have a significant impact on the prediction performance. Although amyloid load saturates several years before symptom onset (Jack et al., 2010b; Yau et al., 2015), several studies show that MCI individuals who are amyloid positive are more likely to convert to dementia in the next 2 to 4 years than those who are amyloid negative (Landau et al., 2012; Jack et al., 2010b; Okello et al., 2009).

The use of EEG or MEG has a significant impact on the performance. However, only 5 experiments using these features were included in the model, it is therefore difficult to conclude if this effect is real, and if it is not due to methodological issues that have not been identified during the quality check.

### 4.3. Longitudinal data

Longitudinal data could give a better view of the evolution of the patient, and hence be more predictive of the progression to AD than cross-sectional data. Nonetheless, we did not find the use of longitudinal data to have a significant effect on the performance. Similar findings are reported in Aksman (2017) for the classification of AD and in Schuster et al. (2015) for progressive diseases in general. Longitudinal analyses are more difficult to design in age-related diseases since there is no temporal marker of disease progression especially before diagnosis. Patients are also seen at different time-points and not all features are acquired at each visit, leading to many missing values. Methodologies for such designs are more exploratory than for cross-sectional approaches (Schiratti et al., 2015; Venkatraghavan et al., 2019)

### 4.4. Algorithm

Table 1 shows that the choice of algorithm has no significant impact on performance. Even if non-linear models seem to be associated to a higher coefficient (0.83) than linear and generalized linear models (−0.47 and 0.19 respectively), these coefficients are far from significant.

The model displayed in Table 2 takes into account the interaction between the model choice and the usage of imaging features. These results show that linear models perform significantly worse than other models (coefficient=-8.11, p=0.001), however the interaction between linear models and imaging features is significantly positive (coefficient = 7.85, p=0.0006); using imaging features therefore leads to a significant increase in performance when using a linear model. Similar conclusions can be drawn from the interaction between generalized linear model and imaging features (coefficient = 4.41, p=0.04), whereas this effect is not significant for non-linear models (coefficient=2.13, p=0.4). By combining the different coefficients, one can see that the best results are obtained using non-linear models. In this case, the use of imaging feature does not significantly impact performance. A possible explanation is that nonlinear models are more powerful and better leverage the information contained in nonimaging data, whereas linear and generalized-linear models have a lower performance on non-imaging data. They therefore benefit from the addition of imaging data, leading to a performance similar to the one obtained using non-linear models.

**Table 2:** Impact of method characteristics, taking into account the interaction between the model type and the use of imaging features. This table shows the coefficients obtained using the linear mixed-effect model described in section 2.4.1 on all experiments, the associated p-values and corrected p-values. The last columns shows the number of experiments using the given characteristic, out of the 120 experiments included in the model. Benjamini-Hochberg procedure was applied to get corrected p-values. coeff.:coefficient of the characteristics in the mixed effect model; APOE: Apolipoprotein E; ADNI: Alzheimer’s Disease Neuroimaging Initiative; NA: not applicable

| Characteristic | coeff. | p-value | corrected p-value | number of exp. |
| --- | --- | --- | --- | --- |
| intercept | 78 | **0** | **0** | NA |
| linear model | -8.1 | **$1*10^{-03}$** | **$5.5*10^{-03}$** | 23 |
| generalized linear model | -3.7 | 0.12 | 0.25 | 28 |
| non linear model | -0.13 | 0.96 | 0.96 | 50 |
| imaging features | -1.6 | 0.42 | 0.52 | 94 |
| cognitive features | 2 | **0.028** | 0.073 | 53 |
| socio-demographic features and APOE | 2.4 | **0.012** | **0.04** | 49 |
| biomarkers | 0.92 | 0.28 | 0.45 | 19 |
| other features | 0.87 | 0.31 | 0.45 | 12 |
| longitudinal | 0.35 | 0.72 | 0.82 | 13 |
| ADNI | -1.42 | 0.26 | 0.45 | 106 |
| number of subjects | -0.066 | 0.96 | 0.96 | NA |
| interaction: linear model and imaging features | 7.85 | **$5.8*10^{-04}$** | **$4.6*10^{-03}$** | 19 |
| interaction: generalized linear model and imaging features | 4.41 | **0.036** | 0.083 | 21 |
| interaction: non linear model and imaging features | 2.13 | 0.41 | 0.52 | 38 |
| individual intercept | 2.27 | **$8.6*10^{-03}$** | **0.034** | NA |

### 4.5. Other methodological characteristics

One could expect the performance to increase when the data set size increases, however we find that the effect of the number of subjects is not significant (coefficient=- 0.39, p=0.76). The impact of data set size is further investigated in 5.1.2.

The impact of using the ADNI data set is not significant (coefficient=0.011, p=0.995). This finding is mitigated by the fact that our results slightly vary when using all experiments or only the ADNI experiment. As only 14 included experiments do not use the ADNI database it is difficult to estimate the impact of its usage independently from the other characteristics.

Although we used a hierarchical grouping of the variables in order to have more statistical power, few p-values and fewer corrected p-values are significant. This small number of significant effects means that the variance of the reported performance measures is high compared to the effect sizes.

## 5. Design of the decision support system and methodological issues

### 5.1. Identified issues

#### 5.1.1. Lack or misuse of test data

The lack of a test data set is observed in 7.3% of experiments. In 16% of articles using feature selection, it is performed on the whole data set, and 8% of articles do not describe this step well enough to draw conclusions. Other data leakage (use of the test set for decision making) is identified in 8% of experiments, and is unclear for 4%.

Overall, 26.5% of articles use the test set in the training process, to train the algorithm, choose the features or tune the parameters. This issue, and in particular performing feature selection on the whole data set, has also been pointed out by Arbabshirani et al. (2017) in the context of brain disorder prediction.

#### 5.1.2. Performance as a function of data set size

We plotted the AUC against the number of individuals for each experiment in Figure 3, with the colored dots representing experiments with identified issues. The colored dots show that there is a higher prevalence of studies with identified issues among high-performance studies: a methodological issue has been identified in 18.5% of experiments with an AUC below 75%, whereas this proportion rises to 36.4% for experiments with an AUC of 75% or higher (significant difference, with p = 0.006).

**Figure 3:**
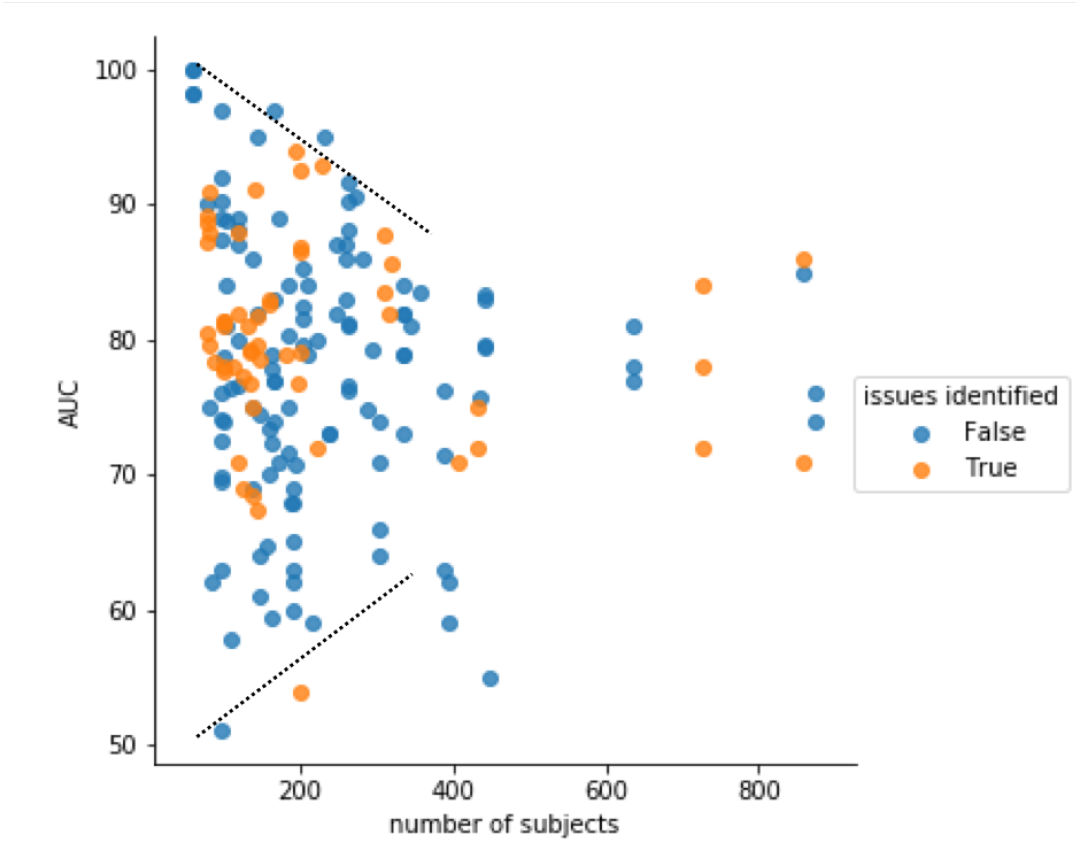
Relationship between the AUC (area under the ROC curve) and the number of individuals. The black dotted lines represent the upper and lower limits.

We can observe an upper-limit (shown in dashed line) decreasing when the number of individuals increases, suggesting that high-performance achieved with a small number of subjects might be due to overfitting. This phenomenon has already been identified by Arbabshirani et al. (2017) and Varoquaux (2018) regarding the use of neuroimaging for brain disorders.

A lower limit is also visible, with the AUC increasing with the number of individuals. This may reflect the fact that, on average, methods generalize better when correctly trained on larger data sets. But it might also suggest that it is harder to publish a method with a relatively low performance if it has been trained on a large number of subjects, such a paper being then considered as reporting a negative result. Within papers also, authors tend to focus on their best performing method, and rarely explain what they learned to achieve this. The machine learning field has the chance to have simple metrics, such as AUC or accuracy, to compare different methods on an objective basis. However, we believe that one should use such metrics wisely not to discourage the publication of innovative methodological works even if it does not yield immediately better prediction performance, and not to overshadow the need to better understand why some methods work better than others.

As the number of subjects increases, the two lines seem to converge to an AUC of about 75%, which might represent the true performance for current state-of-the-art methods.

#### 5.1.3. Use of features of test subjects

Feature embedding is performed on the whole data set in 6.8% of experiments, meaning that the features of the test individuals are used for feature embedding during the training phase. As the diagnosis of the test individuals is often not used for feature embedding, as it is for feature selection, performing it on test individual can be considered a less serious issue than for feature selection. It however requires to re-train the algorithm each time the prediction has to be made on a new individual, which is not suited for a use in clinical practice.

#### 5.1.4. Use of the diagnosis date

In 5.6% of the experiments, the date of AD diagnosis is used to select the input visit of pMCI individuals, for training and testing. As explained in section 2.3, this practice can prevent the generalization of the method to the clinical practice, as the progression date of test individuals is by definition unknown.

This type of experiments answers the question “may one detect some characteristics in the data of a MCI patient 3 years before the diagnosis which, at the same time, is rarely present in stable MCI subjects?”. Which should not be confused with: “can such characteristics predict that a MCI patient will progress to AD within the next 3 years”. What misses to conclude about the predictive ability is to consider the MCI subjects who have the found characteristics and count the proportion of them who will not develop AD within 3 years.

This confusion typically occurred after the publication of Ding et al. (2018). The paper attracted a great attention from general media, including a post on Fox News (Wooller, 2018), stating “Artificial intelligence can predict Alzheimer’s 6 years earlier than medics”. However, the authors state in the paper that “final clinical diagnosis after all follow-up examinations was used as the ground truth label”, thus without any control of the follow-up periods that vary across subjects. Therefore, a patient may be considered as a true negative in this study, namely as a true stable MCI subject, whereas this subject may have been followed for less than 6 years. There is no guarantee that this subject is not in fact a false negative for the prediction of diagnosis at 6 years.

#### 5.1.5. Choice of time-to-prediction

We found that 22.6% of experiments work on separating pMCI from sMCI, regardless of their time to progression to dementia. We advise against this practice, as the temporal horizon at which the individuals are likely to progress is an important information in clinical practice. Methods predicting the exact progression dates, such as what is asked in the Tadpole challenge (Marinescu et al., 2018), should be favored over methods predicting the diagnosis at a given date.

The other experiments have set a specific time to prediction, often between 1 and 3 years, meaning that they intend to predict the diagnosis of the individual at the end of this time interval. Figure 4 shows the evolution of the accuracy of these methods tested on ADNI with respect to the time to prediction. The time to prediction did not have a significant effect on AUC, accuracy, balanced accuracy, specificity nor sensitivity. Figure 4 also shows the accuracy that one would get on ADNI when using a constant prediction, that is predicting that all individuals stay MCI at future time points. The accuracy of this constant prediction has been computed using the proportion of MCI remaining stable at each visit. We show that most methods predicting the progression to AD within a short-term period smaller than 3 years do not perform better than this constant prediction. This finding is consistent with results from the Tadpole challenge (Marinescu et al., 2020), in which no participants significantly outperformed the constant prediction, to which a random noise was added, on prediction of cognitive scores within a 18 month period. We therefore advise to use a time to prediction of at least 3 years. For shorter time intervals the proportion of MCI individuals progressing to AD is so small that predicting that all individuals remain stable gives a better accuracy than most proposed methods.

**Figure 4:**
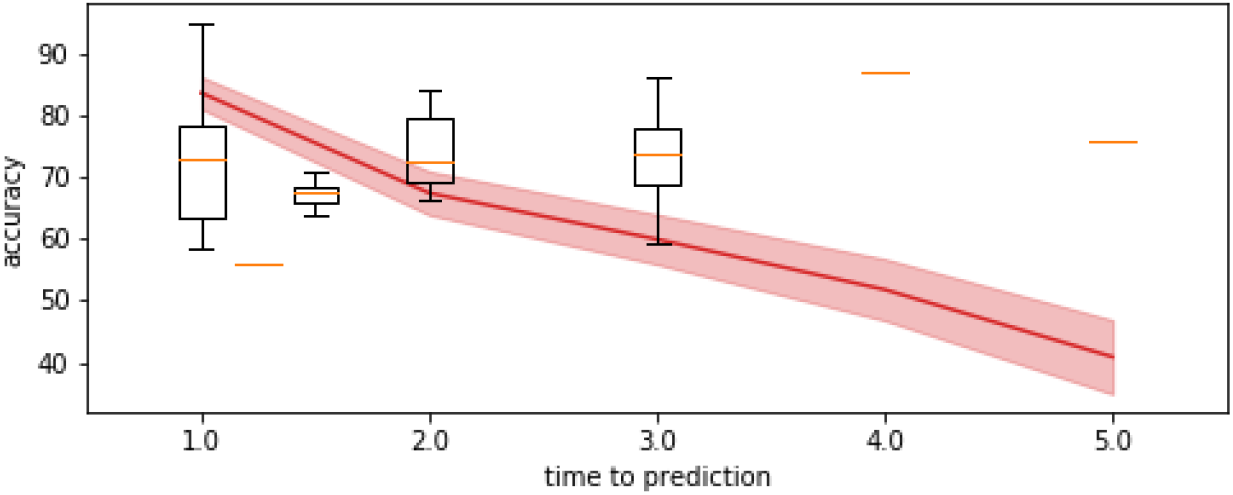
Evolution of the performance with respect to the time to prediction. Box plots represent the accuracy reported in the articles using ADNI included in this review. The straight line represents the accuracy that we computed by predicting that all MCI subjects remain MCI, that is the proportion of MCI subjects in ADNI who remain MCI at the follow-up visit. The shaded area corresponds to the 90% confidence interval. Although some papers in the literature use a sub-set of ADNI and not all ADNI, this plot still shows that results reported in the literature do not out-perform the naive constant prediction for time-to-predictions smaller than 3 years. This comparison is rarely done in the articles.

This fact also shows that the accuracy may be arbitrarily increased by using a cohort with a large proportion of stable subjects. The algorithm may then yield high accuracy by mimicking a constant predictor. This effect may be alleviated by optimizing the balanced accuracy instead of the accuracy.

#### 5.1.6. Problem formulation and data set choice

A common theme that arises from the previous issues is that the methods are not always designed to be the most useful in clinical practice. It is for example true of methods that do not use a specific time-to-prediction, or that use the date of AD diagnosis to select the included visits.

More generally, we think the most useful decision support system should not only focus on Alzheimer’s disease but perform differential diagnosis. Clinicians do not usually need to distinguish between individuals who will develop AD and individuals who will not develop any neurological disorder. They most likely need help to determine which disorder an MCI individual is likely to develop. Unfortunately, no widely available data set allows the development methods for differential diagnosis to date. Methods focusing on AD should therefore target individuals who have already been identified as at risk of developing AD, by providing insight on the date at which this conversion is likely to happen. Such methods could be trained on MCI subjects that are at risk to develop Alzheimer’s disease, defined for instance as the ones who have a MMSE of 27 or smaller and are amyloid positive. In addition to being closer to what can be expected in clinical practice, such data sets of at risk subjects should include a larger proportion of pMCI, leading to a better performance compared to the constant prediction. For example in ADNI, 71.6% of MCI subjects stay stable 2 years after inclusion, whereas this proportion drops to 53.7% for MCI subjects who are amyloid positive and have a MMSE of 27 or lower. Similarly, one should think carefully of the possible other biases introduced by the selection of sMCI or pMCI sub-sets, for instance bias in age, gender or cognitive state. One choice is to match the two sub-sets for these factors. This choice is justified for the detection of the features that are specific to the progressers and to the stable MCI. However, to analyze the performance of a decision support system, one should better reproduce the biases within the population that will be tested by the system in a real case scenario.

The diagnosis of Alzheimer’s disease highly depends on the clinical practice, and varies greatly across sites and countries (Beach et al., 2012). Therefore, the short-term prediction of progression to Alzheimer’s disease is unlikely to generalize well outside the well controlled environment of a research study. Studies on clinical data sets, such as performed in Archetti et al. (2019) regarding the prediction of current diagnosis, could assess how these methods would perform in clinical settings. An interesting alternative may be to predict the changes in the imaging or clinical biomarkers in time rather than the change in diagnosis, such as in Koval et al. (2020), Iddi et al. (2019) and Marinescu et al. (2020).

### 5.2. Need to adhere to best practice guidelines

Given the number of methodological issues that we found in the preparation of this review and that we have discussed above, we feel the need to list here several best practices recommendations.

We first list general guidelines to ensure best generalization of the method and limit the risk for overfitting, following Hastie et al. (2009); Bishop (2006); Géron (2019); Poldrack et al. (2019):

- Separate train and test data sets by using independent cohorts or, if not available, cross-validation. Following Hastie et al. (2009); Varoquaux et al. (2017); Borra and Di Ciaccio (2010); Davison and Hinkley (1997); Kohavi (1995), guidelines for best practices recommend to favor k-fold, repeated k-fold and repeated holdout over leave-one-out method.
- No element of the test data set, both labels and features, should be used except for performance evaluation. In particular, parameter tuning should not be performed on test data, therefore nested cross-validation or train, validation and test splits should be used to tune the algorithm parameters.
- Use a large data set or pool different cohorts to obtain a large data set. Figure 3 shows that overfitting is reduced for more than 300 subjects, at which point the maximum AUC seems to stabilize. This is concordant with results from Arbabshirani et al. (2017), showing a similar point around 200 subjects. Similarly, Poldrack et al. (2019) recommends using data sets of at least several hundred subjects.

We also compile a list of guidelines to carefully design the experiments so that they could support the conclusion about the predictive performance of the method which, in this particular context, includes:

- pre-registration of the time window within which one aims to predict conversion to AD, as we show that performance may greatly vary depending on the timewindow and that no conclusion could be drawn regarding the ability to predict the future without it,
- definition of data sets that best reflect the use of the method in the clinical practice, for instance by selecting subjects that would be considered at risk of developing the disease rather than all possible subjects in ADNI, or by using sex ratio, distribution of age, cognitive state and other similar factors that best mimic the population characteristics that will be tested by the system.
- systematic benchmark of the method against the prediction that the subjects will remain stable over time, as we show that this naive method often outperforms proposed method with a time-to-prediction smaller than 3 years.

## 6. Conclusion

We conducted a systematic and quantitative review on the automatic prediction of the evolution of clinical status of MCI individuals. We reported results from 234 experiments coming from 111 articles. We showed that studies using cognitive variables or FDG PET reported significantly better results than studies that did not, and that including other feature types does not significantly improve performance compared to using cognition or FDG PET alone. These modalities should be further explored, cognition because it can be easily collected in clinical routine, and FDG PET for the interest it might represent for the imaging community and for increasing our understanding of the disease. On the other hand, we showed that using solely T1 MRI yields a significantly lower performance, despite the great number of methods developed for this imaging modality. These findings call into question the role of imaging, and more particularly of MRI, for the prediction of the progression of MCI individuals to dementia. In light of this review, we believe that one should give higher priority to other modalities. More specific cognitive tests could be created, and the impact of using digitized tests, that can be frequently used at home by the patients themselves, should be studied. The creation of digitized tests for clinical routine, such as proposed by Souillard-Mandar et al. (2016); Müller et al. (2017); Schinle et al. (2018) is a first step in this direction.

We identified several key points that should be checked when creating a method which aims at being used as a clinical decision support. When possible, an independent test set should be used to evaluate the performance of the method, otherwise a test set can be separated by carefully splitting the cohort. In any case, the test individuals should not be used to make decisions regarding the method, such as the selection of the features or parameter tuning. The time window in which one aims at predicting the progression to AD should be pre-registered, as the temporal horizon at which an individual is likely to progress to AD is a useful information for clinicians. Alzheimer’s disease being a very slowly progressive disease, algorithm performance should be systematically compared with the prediction that no change will occur in the future. We have shown indeed that the constant prediction may yield very high performance depending on the time frame of the prediction and the composition of the cohort. Finally, the cohort on which the method is tested should be carefully chosen and defined, so as to reflect the future use in clinical practice as best as possible. We noticed that there is often a confusion between two different objectives: understanding the specificities of subjects who will or will not convert to dementia on the one hand, and predicting the progression to dementia on the other hand. Experiments are often designed to address the first objective, but results are then misinterpreted in relation with the second objective. Addressing each objective requires indeed a rather different experimental design.

Following the guidelines will help to design better systems that would eventually lead to similar results in real life. In any case, the final evaluation of such systems will be done in a prospective manner, either in the framework of a challenge like the TADPOLE challenge (Marinescu et al., 2018, 2019, 2020), or even better in a prospective clinical trial (Bruun et al., 2019).

This review focused on the prediction of progression to dementia, as this problem has, by far, attracted most attention from the scientific community. Nevertheless, predicting the future values of the biomarkers or the images may be of greater interest for such clinical decision support systems to be adopted in practice (Marinescu et al., 2020; Koval et al., 2020; Ansart, 2019).

## Data Availability

The data gathered for this study is availbale at https://gitlab.com/icm-institute/aramislab/mci-progressionreview. The issues identified in each article were removed from this open-access table. They can be requested to
the corresponding author.

## Acknowledgements

Federica Cacciamani, Baptiste Couvy-Duchesne, Pascal Lu and Wen Wei participated in reading articles to conduct this review.

We thank the reviewers for their insightful comments that helped us to improve the manuscript, including Gaël Varoquaux who purposely disclosed his name.

The research leading to these results has received funding from the program “Investissements d’avenir” ANR-10-IAIHU-06 (Agence Nationale de la Recherche-10-IA Institut Hospitalo-Universitaire-6) from the European Union H2020 program (project EuroPOND, grant number 666992, project HBP SGA1 grant number 720270), from the ICM Big Brain Theory Program (project DYNAMO, project PredictICD), from the Inria Project Lab Program (project Neuromarkers), from the European Research Council (to Dr Durrleman project LEASP, grant number 678304), from the Abeona Foundation (project Brain@Scale). OC is supported by a “contrat d’interface local” from AP-HP. China Scholarship Council supports J.W’s work on this topic.

Data used in preparation of this article were obtained from the Alzheimer’s Disease Neuroimaging Initiative (ADNI) database (adni.loni.usc.edu). As such, the investigators within the ADNI contributed to the design and implementation of ADNI and/or provided data but did not participate in analysis or writing of this report. A complete listing of ADNI investigators can be found at: http://adni.loni.usc.edu/wp-content/uploads/how_to_apply/ADNI_Acknowledgement_List.pdf

## Appendix A. Supplementary Materials

### Appendix A.1. Query

The full query was:

TITLE-ABS-KEY (“alzheimer’s” OR alzheimer OR ad) AND TITLE-ABS-KEY (“Mild Cognitive Impairment” OR “MCI”) AND TITLE-ABS-KEY ((predicting OR prediction OR predictive) AND (conversion OR decline OR progression OR onset) OR prognosis) AND TITLE-ABS-KEY (accuracy OR roc OR auc OR specificity OR sensitivity) AND (TITLE-ABS-KEY (“Deep learning” OR “ neural network” OR “neural networks” OR “convolutional network” OR “convolutional networks” OR “bayesian network” OR “bayesian networks”) OR TITLE-ABS-KEY (“Matrix completion” OR “Support vector machine” OR “linear mixed-effect” OR “logistic regression” OR “Random Forest” OR “ kernel classifier” OR “kernel” OR “decision tree” OR “ decision trees” OR “least-squares”) OR TITLE-ABS-KEY (“ Machine learning” OR “pattern recognition” OR “pattern classification” OR “classifier” OR “algorithm” OR “ classification”))

### Appendix A.2. Selection process diagram

The process used to select the articles included in the review is shown in Figure S1.

### Appendix A.3. Reported items

For each article, the following elements were reported:

- number of MCI subjects progressing to AD;
- number of stable MCI subjects;
- time to prediction;
- used cohorts;
- use of socio-demographic features (yes/no);
- use of APOE (yes/no);
- use of general cognitive features (yes/no);
- use of domain-targeted cognitive features (yes/no);
- use of new, home-made cognitive features (yes/no);
- use of voxel based features from T1 MRI (yes/no);
- use of regions of interest on the whole brain, from T1 MRI (yes/no);
- use of selected regions of interest from T1 MRI (yes/no);
- use of white matter hyper-intensities (yes/no);
- use of PET FDG features (yes/no);
- use of PET amyloid features (yes/no);
- use of PET tau features (yes/no);
- use of CSF features (yes/no);
- use of amyloid status (yes/no);
- use of DTI features (yes/no);
- use of functional MRI features (yes/no);
- use of EEG or MEG features (yes/no);
- use of other features (yes/no, precision given as a free note);
- use of longitudinal features (yes/no);
- is feature selection performed (yes/no);
- used algorithm (categories defined below);
- validation method (categories defined bellow);
- feature selection performed on the whole data set (yes/no/unclear);
- feature embedding performed on the whole data set (yes/no/unclear);
- selection of the input visit of the test subjects using their date of progression to AD (yes/no);
- other data leakage (use of the test set to make decisions) (yes/no/unclear);
- other issue (yes/no)
- AUC value;
- accuracy value;
- balanced accuracy value;
- sensitivity value;
- specificity value;

**Figure S1:**
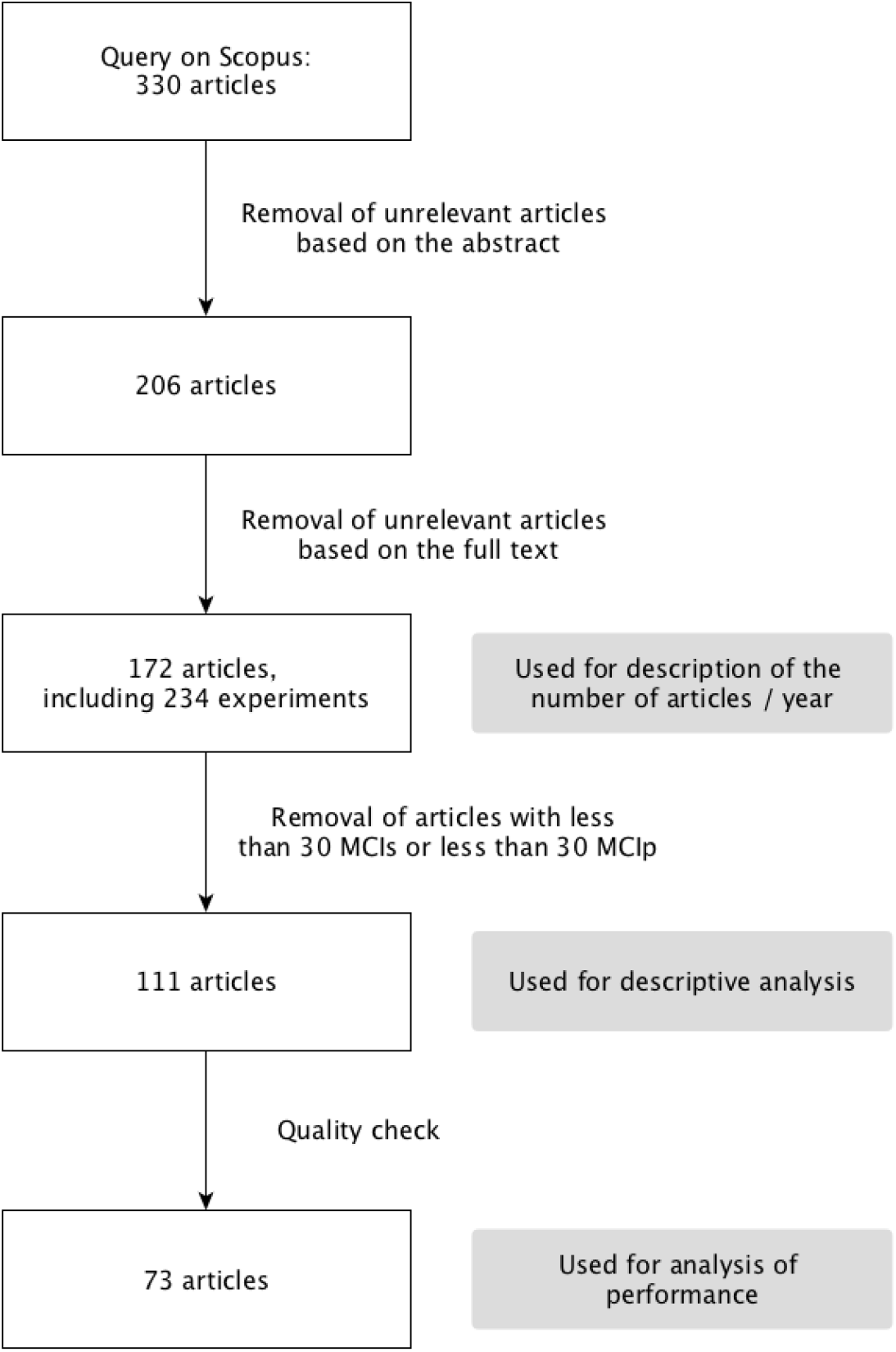
Diagram representing who the articles were selected

Free notes describing the issues, or important points that did not fit in the previous list, were added.

The possible algorithm categories were added by the readers and aggregated. The final list was: bayesian algorithms, classification by clinicians, gaussian process, linear discriminant analysis (LDA), low rank matrix completion (LRMC), linear regression, logistic regression, manifold learning, multiple kernel learning, neural network, orthogonal partial least square (OPLS), random forest, regularized logistic regression, linear support vector machine (linear SVM), non-linear SVM, SVM with unknown kernel (simply noted as SVM), survival analysis, use of a threshold and others (including home-made algorithms).

The same process was used to create the cross-validation category list, composed of: 10-fold, k-fold, repeated k-fold, leave one out, out of the bag, single split, repeated single split, validation on an independent cohort, validation on different groups (when the algorithm is trained on separating AD and CN subjects, and tested on predicting the progression of MCI subjects), none, not described (when the use of cross-validation is mentioned but the used validation method is not described) and not needed (for thresholding with a manually chosen threshold for example).

### Appendix A.4. Additional performance analysis Appendix

#### A.4.1. On all experiments

As explained in 2.4.1, we first built a coarse model, grouping the characteristics into broad categories. The results are shown in Table S1. As several of the broad categories had a significant effect on performance (imaging and cognitive features), these categories were expanded into finer ones, building the model shown in Table S2. The significant categories were expanded once again (PET and socio-demographic features and APOE, EEG/MEG was already expanded at maximum level), leading to the model shown in table 1. As all the significant characteristics were expanded at the maximum level, no further model was created.

#### Appendix A.4.2. On the ADNI experiments

We also analysed the impact of method characteristic on performance using only the experiments performed on ADNI. Table S3 shows the model obtained by using the broadest characteristic categories. Table S4 shows the results obtained by expanding the categories significant in Table S3 (imaging and cognitive features). Table S5 shows the results obtained by expanding the categories significant in Table S4 (PET and T1 features, domain targeted cognitive features being expanded at maximum level).

**Table S1:**
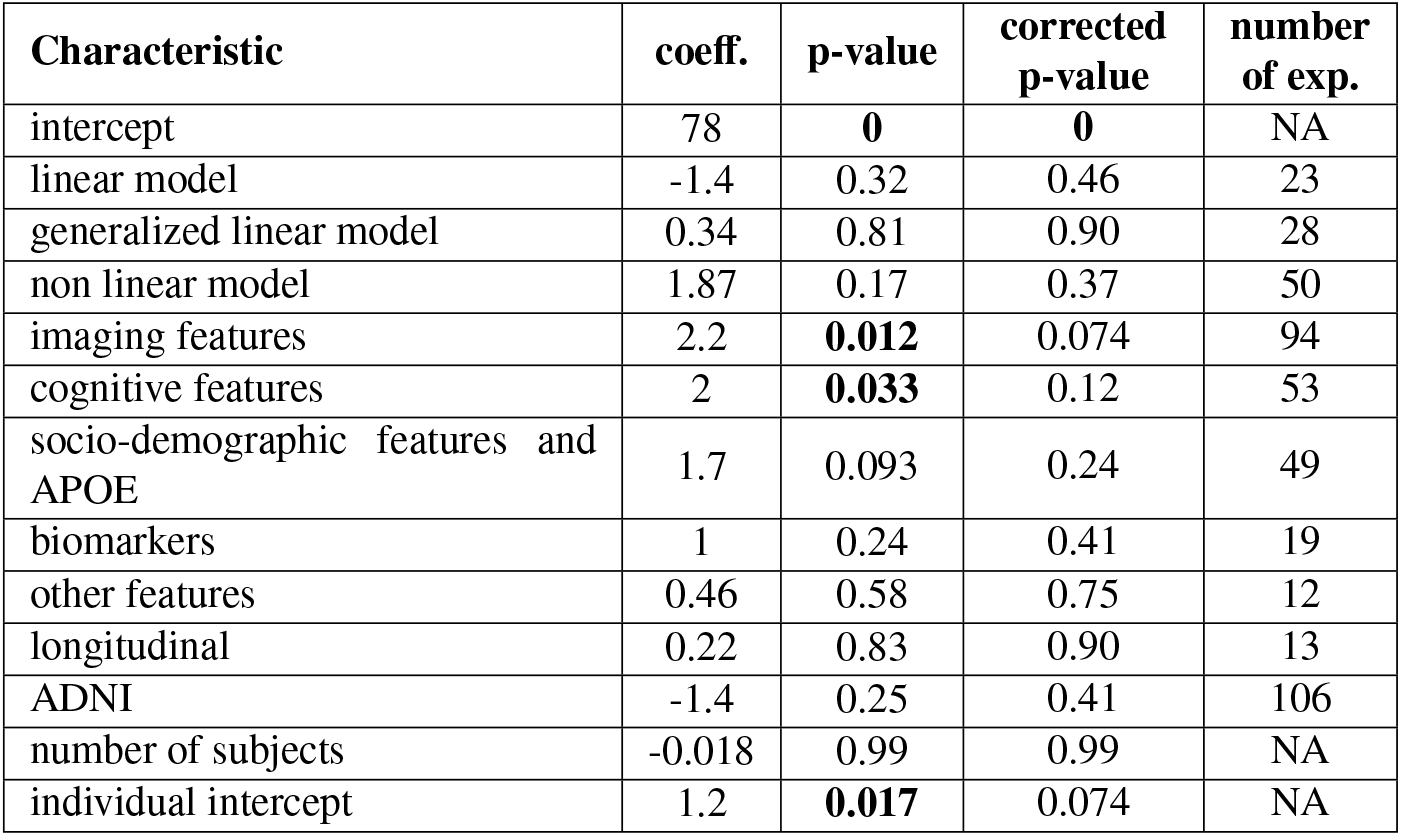
Impact of method characteristics on all experiments, using the broadest categories. This table shows the coefficients obtained using the linear mixed-effect model described in section 2.4.1 on all experiments, the associated p-values and corrected p-values. The last columns shows the number of experiments using the given characteristic, out of the 120 experiments included in the model. Benjamini-Hochberg procedure was applied to get corrected p-values. coeff.:coefficient of the characteristics in the mixed effect model; APOE: Apolipoprotein E; ADNI: Alzheimer’s Disease Neuroimaging Initiative; NA: not applicable

| Characteristic | coeff. | p-value | corrected p-value | number of exp. |
| --- | --- | --- | --- | --- |
| intercept | 78 | **0** | **0** | NA |
| linear model | -1.4 | 0.32 | 0.46 | 23 |
| generalized linear model | 0.34 | 0.81 | 0.90 | 28 |
| non linear model | 1.87 | 0.17 | 0.37 | 50 |
| imaging features | 2.2 | **0.012** | 0.074 | 94 |
| cognitive features | 2 | **0.033** | 0.12 | 53 |
| socio-demographic features and APOE | 1.7 | 0.093 | 0.24 | 49 |
| biomarkers | 1 | 0.24 | 0.41 | 19 |
| other features | 0.46 | 0.58 | 0.75 | 12 |
| longitudinal | 0.22 | 0.83 | 0.90 | 13 |
| ADNI | -1.4 | 0.25 | 0.41 | 106 |
| number of subjects | -0.018 | 0.99 | 0.99 | NA |
| individual intercept | 1.2 | **0.017** | 0.074 | NA |

**Table S2:** Impact of method characteristics on all experiments, after refining the categories that were significant in table S1. This table shows the coefficients obtained using the linear mixed-effect model described in section 2.4.1 on all experiments, the associated p-values and corrected p-values. The last columns shows the number of experiments using the given characteristic, out of the 120 experiments included in the model. Benjamini-Hochberg procedure was applied to get corrected p-values. coeff.:coefficient of the characteristics in the mixed effect model; PET: positron emission tomography; EEG: electroencephalography; MEG: magnetoencephalography; APOE: Apolipoprotein E; ADNI: Alzheimer’s Disease Neuroimaging Initiative; NA: not applicable

| Characteristic | coeff. | p-value | corrected p-value | number of exp. |
| --- | --- | --- | --- | --- |
| intercept | 78 | **0** | **0** | NA |
| linear model | -0.79 | 0.67 | 0.86 | 23 |
| generalized linear model | 0.2 | 0.89 | 0.92 | 28 |
| non linear model | 0.95 | 0.49 | 0.73 | 50 |
| T1 features | 0.99 | 0.21 | 0.53 | 77 |
| PET | 2.82 | **$6.8 \times 10^{-03}$** | **0.03** | 25 |
| white matter hyper-intensities | -0.64 | 0.44 | 0.72 | 3 |
| EEG/MEG | 3.5 | **$1.1 \times 10^{-03}$** | **0.01** | 5 |
| general cognitive features | 0.5 | 0.67 | 0.86 | 49 |
| domain targeted cognitive features | 2.2 | 0.051 | 0.15 | 25 |
| new or specific cognitive features | 1.1 | 0.4 | 0.72 | 2 |
| socio-demographic features and APOE | 2.8 | **$4.7 \times 10^{-03}$** | 0.028 | 49 |
| biomarkers | 0.84 | 0.32 | 0.72 | 19 |
| other features | 0.75 | 0.37 | 0.72 | 12 |
| longitudinal | 0.34 | 0.74 | 0.88 | 13 |
| ADNI | 0.2 | 0.9 | 0.92 | 106 |
| number of subjects | -0.13 | 0.92 | 0.92 | NA |
| individual intercept | 2.7 | **0.03** | 0.11 | NA |

**Table S3:** Impact of method characteristics on ADNI experiments, using the broadest categories. This table shows the coefficients obtained using the linear mixed-effect model described in section 2.4.1 on the ADNI experiments, the associated p-values and corrected p-values. The last columns shows the number of experiments using the given characteristic, out of the 106 experiments included in the model. Benjamini-Hochberg procedure was applied to get corrected p-values. coeff.:coefficient of the characteristics in the mixed effect model; APOE: Apolipoprotein E; NA: not applicable; ADNI: Alzheimer’s Disease Neuroimaging Initiative

| Characteristic | coeff. | p-value | corrected p-value | number of exp. |
| --- | --- | --- | --- | --- |
| intercept | 78 | **0** | **0** | NA |
| linear model | 0.5 | 0.72 | 0.86 | 16 |
| generalized linear mode | 0.91 | 0.51 | 0.68 | 27 |
| non linear mode | 1.7 | 0.19 | 0.45 | 48 |
| imaging features | 2.3 | **$3.5 \times 10^{-04}$** | **$2.1 \times 10^{-03}$** | 87 |
| cognitive features | 2.4 | **$3 \times 10^{-03}$** | **0.012** | 48 |
| socio-demographic features and APOE | 0.099 | 0.92 | 0.96 | 42 |
| biomarkers | 0.6 | 0.46 | 0.68 | 16 |
| other features | 0.79 | 0.26 | 0.52 | 9 |
| longitudinal | 0.86 | 0.34 | 0.59 | 13 |
| number of subjects | 0.074 | 0.96 | 0.96 | NA |
| individual intercept | 4.1 | **$8.9 \times 10^{-03}$** | **0.027** | NA |

**Table S4:** Impact of method characteristics on ADNI experiments, after refining the categories that were significant in table S3. This table shows the coefficients obtained using the linear mixed-effect model described in section 2.4.1 on the ADNI experiments, the associated p-values and corrected p-values. The last columns shows the number of experiments using the given characteristic, out of the 106 experiments included in the model. Benjamini-Hochberg procedure was applied to get corrected p-values. coeff.:coefficient of the characteristics in the mixed effect model; PET: positron emission tomography; APOE: Apolipoprotein E; NA: not applicable; ADNI: Alzheimer’s Disease Neuroimaging Initiative

| Characteristic | coeff. | p-value | corrected p-value | number of exp. |
| --- | --- | --- | --- | --- |
| intercept | 77 | **0** | **0** | NA |
| linear model | 1.9 | 0.11 | 0.25 | 16 |
| generalized linear model | 1.4 | 0.26 | 0.4 | 27 |
| non linear model | 1.3 | 0.23 | 0.4 | 48 |
| T1 features | 1.4 | **$6.3 \times 10^{-03}$** | **0.022** | 77 |
| PET | 4.6 | **$3.8 \times 10^{-09}$** | **$2.7 \times 10^{-08}$** | 25 |
| general cognitive features | 0.64 | 0.48 | 0.56 | 46 |
| domain targeted cognitive features | 2.3 | **$9.1 \times 10^{-03}$** | **0.026** | 23 |
| socio-demographic features and APOE | 0.69 | 0.43 | 0.53 | 42 |
| biomarkers | 0.18 | 0.8 | 0.8 | 16 |
| other features | 0.55 | 0.37 | 0.51 | 9 |
| longitudinal | 0.94 | 0.22 | 0.4 | 13 |
| number of subjects | 0.73 | 0.62 | 0.67 | NA |
| individual intercept | 9.9 | **$4.7 \times 10^{-03}$** | **0.022** | NA |

**Table S5:**
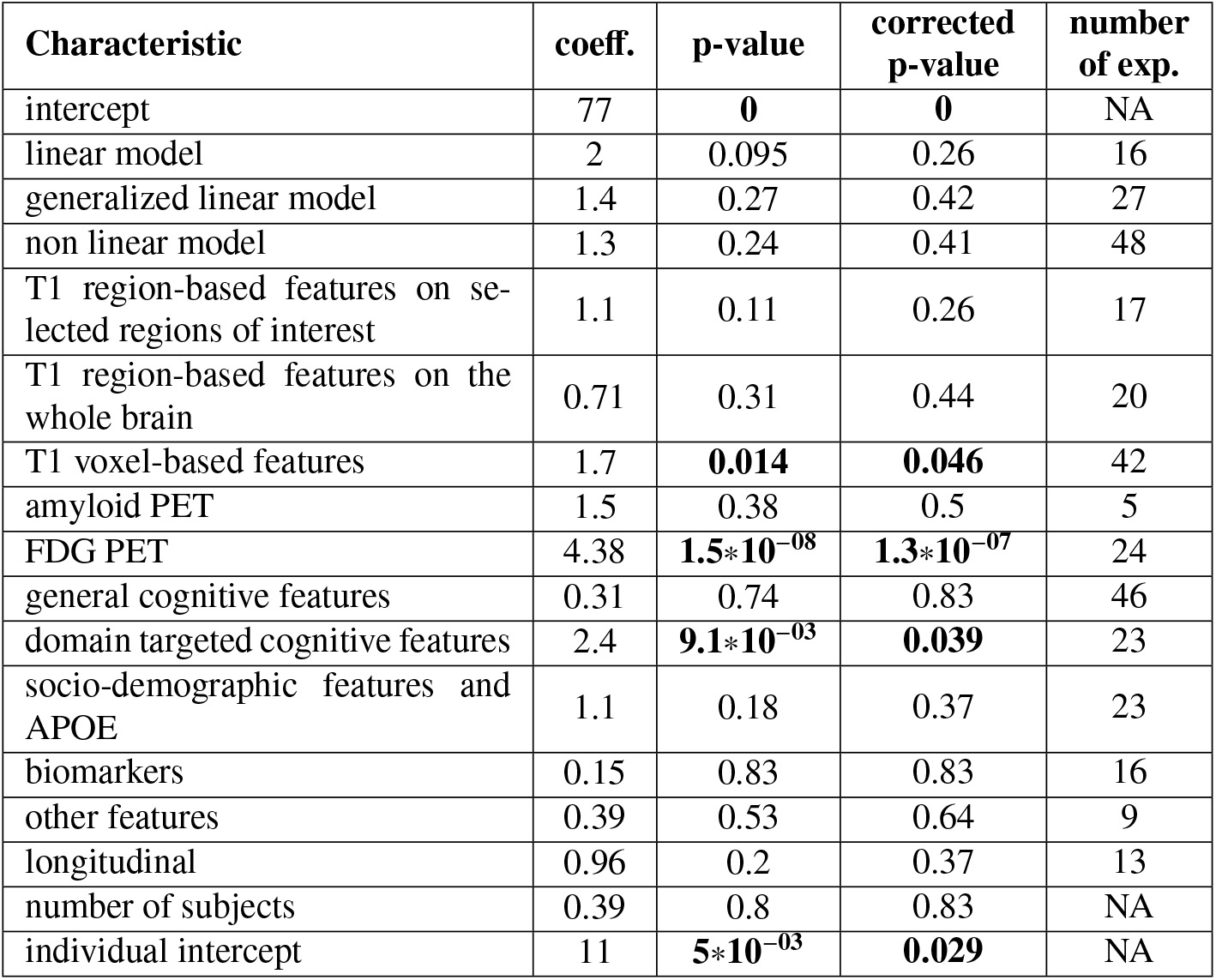
Impact of method characteristics on ADNI experiments, after refining the categories that were significant in table S4. This table shows the coefficients obtained using the linear mixed-effect model described in section 2.4.1 on the ADNI experiments, the associated p-values and corrected p-values. The last columns shows the number of experiments using the given characteristic, out of the 106 experiments included in the model. Benjamini-Hochberg procedure was applied to get corrected p-values. coeff.:coefficient of the characteristics in the mixed effect model; PET: positron emission tomography; FDG: fluorodeoxyglucose; APOE: Apolipoprotein E; NA: not applicable; ADNI: Alzheimer’s Disease Neuroimaging Initiative

### Appendix A.5. Journals and conference proceedings

Table S6 shows the journals and conference proceedings in which more than one included article has been published, and the associated number of articles.

**Table S6:** Number of included articles published in each journal or conference proceedings. Only the journals with more than one included article are shown here. The articles taken into account are the one considered for analysis, and that use a large enough data set.

| **Journal or conference proceedings** | **Number of included articles** |
| --- | --- |
| Journal of Alzheimer's Disease | 12 |
| NeuroImage | 11 |
| Lecture Notes in Computer Science | 7 |
| PLoS ONE | 9 |
| Neurobiology of Aging | 6 |
| Neurology | 3 |
| Brain Topography | 3 |
| Current Alzheimer Research | 3 |
| Medical Image Analysis | 3 |
| Frontiers in Aging Neuroscience | 3 |
| Scientific Reports | 2 |
| Frontiers in Neuroscience | 2 |
| IEEE Journal of Biomedical and Health Informatics | 2 |
| IEEE Transactions on Biomedical Engineering | 2 |
| NeuroImage: Clinical | 2 |
| Journal of Neuroscience Methods | 2 |

## Appendix B. Articles included in the review

